# Response of Wastewater-Based Epidemiology Predictor for the Second Wave of COVID-19 in Ahmedabad, India: A Long-term Data Perspective

**DOI:** 10.1101/2022.05.28.22275432

**Authors:** Manish Kumar, Madhvi Joshi, Guangming Jiang, Rintaro Yamada, Ryo Honda, Vaibhav Srivastava, Jürgen Mahlknecht, Damia Barcelo, Sabarathinam Chidambram, Anwar Khursheed, David Graham, Chaitanya Joshi

## Abstract

Wastewater-based epidemiology (WBE) monitoring can play a key role in managing future pandemics because it covers both pre-symptomatic and asymptomatic cases, especially in densely populated areas with limited community health care. In the present work, wastewater monitoring was employed in Ahmedabad, India, after the successful containment of the first wave of COVID-19 to predict resurgence of the disease in the expected second wave of the pandemic. Here we show wastewater levels of COVID-19 virus particles (i.e., SARS-CoV-2) positively correlated with the number of confirmed clinical cases during the first wave, and provided early detection of COVID-19 presence before the second wave in Ahmedabad and an WBE-based city zonation plan was developed for health protection. A eight-month data of Surveillance of Wastewater for Early Epidemic Prediction (SWEEP) was gathered, including weekly SARS-CoV-2 RNA wastewater analysis (n=287) from nine locations between September 2020 and April 2021. Across this period, 258 out of 287 samples were positive for least two out of three SARS-CoV-2 genes (N, ORF 1ab, and S). Monitoring showed a substantial decline in all three gene markers between October and September 2020, followed by an abrupt increase in November 2020. Similar changes were seen in March 2021, which preceded the second COVID-19 wave. Measured wastewater ORF-1ab gene copies ranged from 6.1 × 10^2^ (October, 2020) to 1.4 × 10^4^ (November, 2020) copies/mL, and wastewater gene levels typically lead confirmed cases by one to two weeks. The study highlights the value of WBE as a monitoring tool to predict waves within a pandemic, identifying local disease hotspots within a city and guiding rapid management interventions.

**Highlights:**

- Eight-months of SARS-CoV-2 gene variations explicitly predicts 2^nd^ COVID-19 wave.
- 258 out of 287 wastewater samples were positive for SARS-CoV-2 genes.
- WBE offers a lead time of 1-2 weeks relative to clinical cases.
- Model suggests that ORF 1ab gene is the most effective as a marker gene in WBE study.
- WBE RT-PCR screening for pathogens should be mandatory for global health monitoring.

## 1. Introduction

The global pandemic caused due to the severe acute respiratory syndrome coronavirus 2 (SARS-CoV-2), has infected 43 million people across India by April 16^th^, 2022 (WHO, 2022). Cases with mild or no symptoms are often overlooked, leading to inaccuracy in epidemiological models and assessment of disease prevalence. A large number of asymptomatic patients employed a never seen challenges over the verified estimation of disease spread based on clinical surveillance (Rimoldi et al., 2020; Medema et al., 2020). According to earlier studies, 18-45% of patients infected with COVID-19 are asymptomatic, but they are capable of spreading the disease, thereby adversely affecting the actual containment of the disease (Lavezzo et al., 2020; Yang et al., 2020; Mizumoto et al., 2020; Nishiura et al., 2020). Wastewater-based epidemiology (WBE) monitoring has gained tremendous recognition as a viable option for COVID-19 monitoring since nearly 67% of infected people showed the presence of SARS-CoV-2 RNA in feces (Chan et al., 2020; Cheung et al., 2020; Parasa et al., 2020; Wong et al., 2020). COVID patients may shed viruses to wastewater through sputum and saliva (Li et al., 2022). It can be used to detect the arrival and subsequent decline of pathogens as well as provide an early warning of the forthcoming prevalence of the disease within a community (Hata et al., 2021; Kumar et al., 2021a, b). There are certain advantages of using WBE over clinical testing which includes reduced analytic costs. In addition, wastewater contains viruses shed from a large number of people and thus requires far fewer number of samples and less labour than clinical testing to know the presence of infected persons in particular location. However, WBE is less sensitive towards detection of SARS-CoV-2 in comparison to norovirus, probably due to its enveloped nature and low SARS-CoV-2 load in the patient’s fecal matter and other shedding sources (Hata et al., 2020). Further, it is essential to explore the correlation between the SARS-CoV-2 genetic load in wastewater and the number of cases at the district level in each country to evaluate WBE’s potential as an early prediction tool for COVID-19 pandemic.

Even though the capability of WBE monitoring to detect RNA of SARS-CoV-2 has been proven, several constraints and bottlenecks exist towards its practical applicability (Zhu et al., 2021, Tran et al., 2020). It is extremely necessary to match the time-series data of SARS-CoV-2 RNA concentration in the wastewater with the actual clinical survey data in order to confirm the utility and predictability of wastewater monitoring. This is also essential for the adaptation of the *Surveillance of Wastewater for Early Epidemic Prediction (SWEEP)* on the policy level, which has been suspended for various reasons in major parts of the globe (Tiwari et al., 2021). The effectiveness of WBE has been debated actively on the basis of watersheds, catchment type, complexity of sewer systems, and population. If cases reported from the given city have been substantially high, it is pertinent to check the efficacy of SWEEP on the urban scale. Under this framework, four major directions in the domain of SWEEP may be compiled as i) validating the data to unravel the early warning capability of wastewater monitoring for COVID-19 through temporal studies on SARS-CoV-2 RNA detection; ii) need for an increase of WBE monitoring in various parts of the world to generate data from all the levels of COVID-19 situation; iii) developing the model that can utilize Ct-value acquired through SWEEP into significant predictions for effectual COVID-19 pandemic preparedness; and iv) collectively reach to the comprehension of crucial issues like removal, discharge, decay, dilution, and infectivity due to the presence of SARS-CoV-2 RNA in wastewater (Kumar et al., 2021a ; Prevost et al., 2015).

Taking this into consideration, the present study aims to present the wastewater monitoring results from Ahmedabad, India, and its association with the second wave of COVID-19 pandemic by drawing a comparison between the detected concentration of SARS-CoV-2 RNA in wastewater of various parts of the city and the COVID-19 confirmed clinical cases. Clinical surveillance of COVID-19 is often inadequate to classify the city into specific zones based on the requirement of more tests or attention. This is particularly true for poorly resourced regions where a lower number of confirmed cases may be linked to underreporting. In such cases, SWEEP-based information may prove to be critical in zonation of the city and locating hotspots on a city scale. The concentration of SARS-CoV-2 RNA detected in wastewater would shed light on the true prevalence of COVID-19 infection in the sewer catchment, whereas the numbers reported from the clinically reported cases only account for the diagnosed patients thereby excluding the undiagnosed or asymptomatic patients from the process. In this study, we analysed SARS-CoV-2 RNA in the wastewater samples (*n*=287) from 9 different locations, including wastewater pumping stations and sewage treatment plant (STP) of Ahmedabad, India, from September 3rd to April 12th, 2021 (thirty-two weeks). The main objectives of the study were: **a)** to evaluate the implementation of WBE for the prediction of the second wave of COVID-19 in Ahmedabad; **b)** weekly resolution of the SARS-CoV-2 RNA data for eight months in wastewater samples; and **c)** explicating the potential of WBE for identifying hotspots and public health monitoring at the city level.

## 2. Materials and Methods

### 2.1. Study area

Ahmedabad is the seventh largest city in India and the second biggest trade centre in the western part of India, with an estimated population of 8.25 million in 2020 (UN world urbanization prospects 2018). It has a sewage network of 2500 km along with 9 sewage treatment plants (STPs) and 45 sewage pumping stations (SPSs). The existing treatment capacity of the wastewater treatment plant in the city is 990 MLD (MoHUA, 2021).

### 2.2. Sampling approach

The sampling locations were determined by following the approach of Kumar et al. (2021) for the same study area (Ahmedabad). A total of 287 samples from nine different sites of Ahmedabad were analysed weekly to detect SARS-CoV-2 RNA in wastewater. Grab sampling method was used to collect the samples in 250 ml sterile bottles (Tarsons, PP Autoclavable, Wide Mouth Bottle, Cat No. 582240, India). In order to detect any contamination during the transport, we examined blanks in the same type of bottle. The samples were transported and maintained at cooling condition in an ice-box until further process. The samples were processed on the same day after bringing them to the laboratory. All the analyses were conducted in Gujarat Biotechnology Research Centre (GBRC), a laboratory approved by the Indian Council of Medical Research (ICMR), New Delhi.

### 2.3. SARS-CoV-2 gene detection

#### 2.3.1. Precipitation of viral particles

Firstly, 30 mL of each sample was centrifuged at 4000×g (Model: Sorvall ST 40R, Thermo Scientific) in a 50 mL sterile falcon tube for 40 minutes, followed by filtration of supernatant using 0.22-micron syringe filter (Mixed cellulose esters syringe filter, Himedia). After filtrating 25 mL of the supernatant, 2 g of PEG 9000 and 0.437 g of NaCl (17.5 g/L) were mixed in the filtrate, which was incubated at 17°C, 100 rpm overnight (Model: Incu-Shaker™ 10LR, Benchmark). The following day, the mixture was centrifuged at 14000×g (Model: Kubota 6500, Kubota Corporation) for a period of about 90 minutes. After centrifugation, the supernatant was discarded, and the pellet was resuspended in 300µL RNase-free water. The concentrated sample was stored in 1.5ml eppendorf at -40 °C, and this was subsequently used as a sample for RNA isolation.

#### 2.3.2. RNA isolation, RT-PCR and gene copy estimation

NucleoSpin® RNA Virus (Macherey-Nagel GmbH & Co. KG, Germany) isolation kit was used to perform RNA isolation from the pellet with the concentrated virus. MS2 phage provided by TaqPath™ COVID-19 RT-PCR Kit was utilized as an internal control. Some other particulars include **a)** the nucleic acid extraction performed by NucleoSpin® RNA Virus Kit (Macherey-Nagel GmbH & Co. KG, Germany) and Qubit 4 Fluorometer (Invitrogen) was used to estimate RNA concentrations, **b)** evaluation of molecular process inhibition control (MPC) was done through MS2 phage for QC/QA analyses of nucleic acid extraction and PCR inhibition (Haramoto et al., 2018). The methodology has been described in author’s previous works (Kumar et al., 2021 and 2020a). The steps were carried out according to the instructions provided in the Macherey-Nagel GmbH & Co. KG product manual, and RNAs were detected using real-time PCR (RT-PCR).

The detection of SARS-CoV-2 was performed by using TaqMan-based chemistry on Applied Biosystems 7500 Fast Dx Real-Time PCR Instrument (version 2.19 software). For each run, a template of 7 µl of extracted RNA was used with TaqPath™ 1 Step Multiplex Master Mix (Thermofischer Scientific, USA). The final reaction mixture (25 µL) consisted of nuclease-free water (10.50 µL), Master Mix (6.25 µL) and COVID-19 Real-Time PCR Assay Multiplex (1.25 µL). Positive control (TaqPath™ COVID 19 Control), negative control (from extraction run spiked with MS2), and no template control (NTC) were run with each batch. 40 cycles of amplification were set, and results were explained on the basis of the Ct values for three target genes i.e., ORF1ab, N (Nucleocapsid), and S (Spike) proteins of SARS-CoV-2 along with that of MS2 used as an internal control.

Results were considered conclusive/ positive only if two or more genes are detected in the samples. Effective genome concentration was computed semi-qualitatively using the equivalence of 500 copies of SARS-CoV-2 genes as 26 Ct-value (provided with the kit). After this, the RNA amount used as a template and the enrichment factor of wastewater samples during the experimentation were multiplied by the result.

#### 2.3.3. Method of Spearman’s rank correlation coefficient

Since the relationship between the SARS-CoV-2 gene concentration in wastewater and the number of new daily confirmed clinical cases is considered to be non-linear, the relationship was evaluated by Spearman’s rank correlation coefficient. Clinical information is based on data from Ahmedabad City (<u>COVID-19 India</u>), and the number of new daily clinical cases for a total of seven days (the reference date and three days before and after that day) was used to analyse the rank correlation coefficient with the concentration of the SARS-CoV-2 gene in wastewater (to be precise, the virus concentration was substituted by the Ct value). This is because the number of new daily clinical cases is affected by day-of-week variations in the number of tests, including PCR and antigen examination, etc. Specifically, the sampling date was used as the reference, and that reference date was shifted back and forth from the sampling date to analyze the respective rank correlation coefficients. The time lag between the increase or decrease of the SARS-CoV-2 gene concentration (Ct value) in wastewater and that of the number of new daily clinical cases was estimated from the gap between the reference date and sampling date when the rank correlation coefficient was the highest. (A negative time lag indicates that wastewater is detecting trends in the viral infection status faster than the clinical tests.)

## 3. Results and discussion

Variations in SARS-CoV-2 RNA were detected and quantified from influent wastewater samples for eight months (September 2020 to April 2021) to understand the pandemic situation in Ahmedabad, Gujarat, India. Out of 287 samples analyzed in the study, 258 were found positive, comprising two out of three target genes **(Table 1)**. Whereas, 253 samples displayed positive RT-PCR results for each N, ORF 1b, and S genes. The average Ct values for S, N, and ORF 1ab genes were 32.89, 31.84, and 32.48, respectively. The average Ct value of internal control (MS2 bacteriophage was 27.42. Also, no SARS-CoV-2 gene was detected in the negative control samples.

**Table 1.**
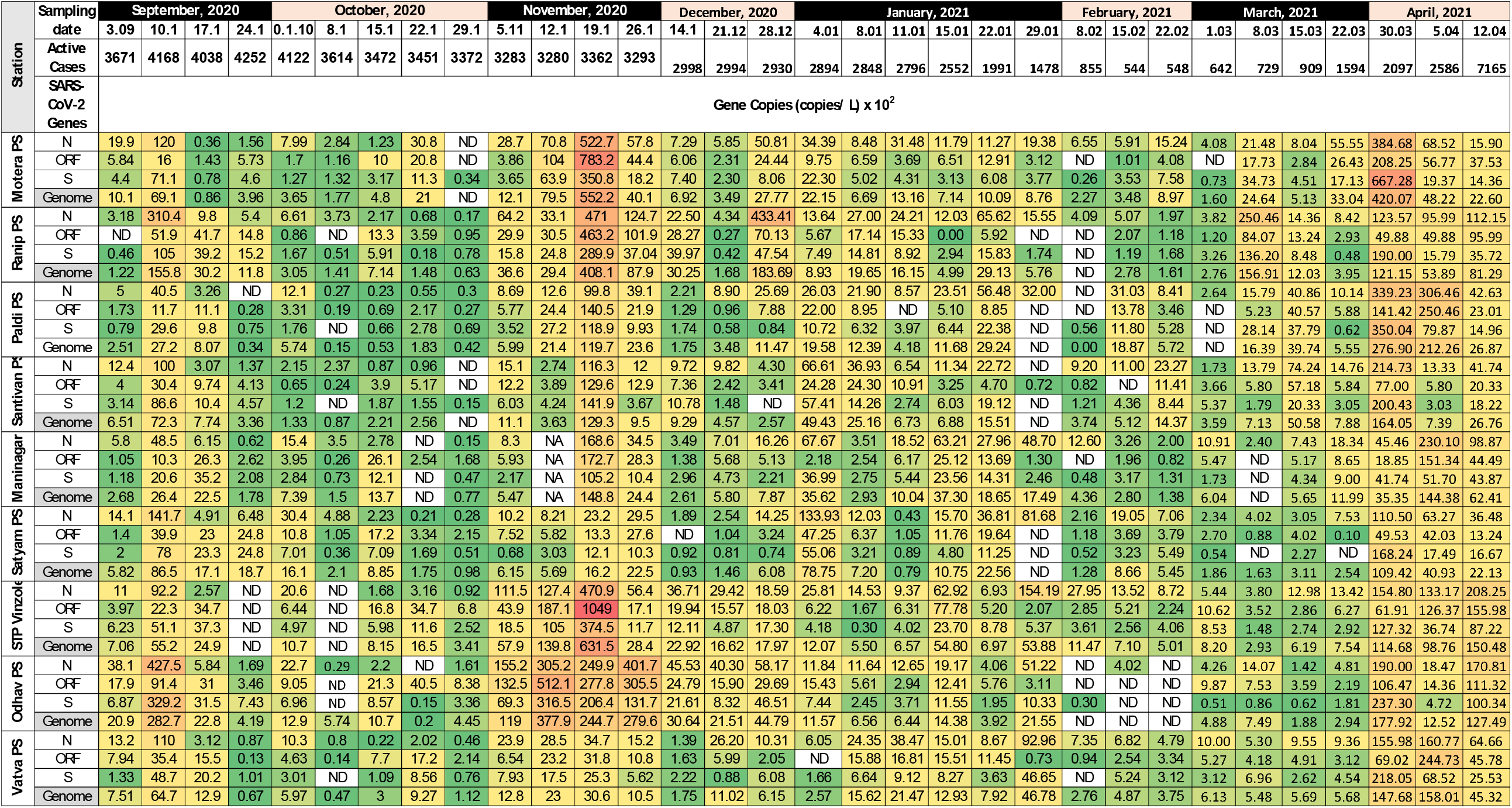
Temporal variation in SARS-CoV-2 genetic material loading found in the influent and effluent samples collected from different wastewater treatment plants

### 3.1. Monthly variation

Monthly variation portrayed a notable decline of 89.7%, 63.7%, and 90.1% in N, ORF-1ab, and S gene concentration (copies/L), respectively, in October compared to September 2020. This was followed by a sharp increase in November 2020 i.e., about 25, 22 and 26 folds in N, ORF 1ab, and S gene, respectively. The concentration of all the three genes started decreasing in the month of December and continued till February 2021. After this, there was a pronounced increase in the concentration of all three genes in the month of March 2021 as compared to February 2021 i.e. about 19, 10, and 6 folds in S, ORF1ab, and N gene, respectively. In April, the average gene concentrations for N, ORF-1ab, and S genes were 10.5×10^3^, 8.3×10^3^, and 3.6×10^3^, respectively. The highly infectious and fatal Delta variation (B.1.617.2), which caused the second wave in India, is responsible for the dramatic increase in gene concentration in March and April.

The descending order of monthly variation in ORF1ab gene concentration in wastewater samples was November> April> March> September> December> January> October> February. Likewise, the decreasing order of N gene in wastewater samples followed a similar pattern and was found in the order of November> April> March> September> December> January> February> October and that of S gene was found to be November> March> April> September> January> December> February> October **(Fig 1 a-d)**. The genome concentration of SARS-CoV-2 RNA was maximum in the month of November (1.1×10^4^ copies/ L), followed by April (7.5×10^3^ copies/L), March (4.5×10^3^ 524 copies/L), September (3.0×10^3^ copies/ L), December (1.8×10^3^ copies/ L), January (1.6×10^3^ copies/ L), February (4.7×10^2^ copies/ L) and October (4.5×10^2^ copies/L).

**Fig. 1.**
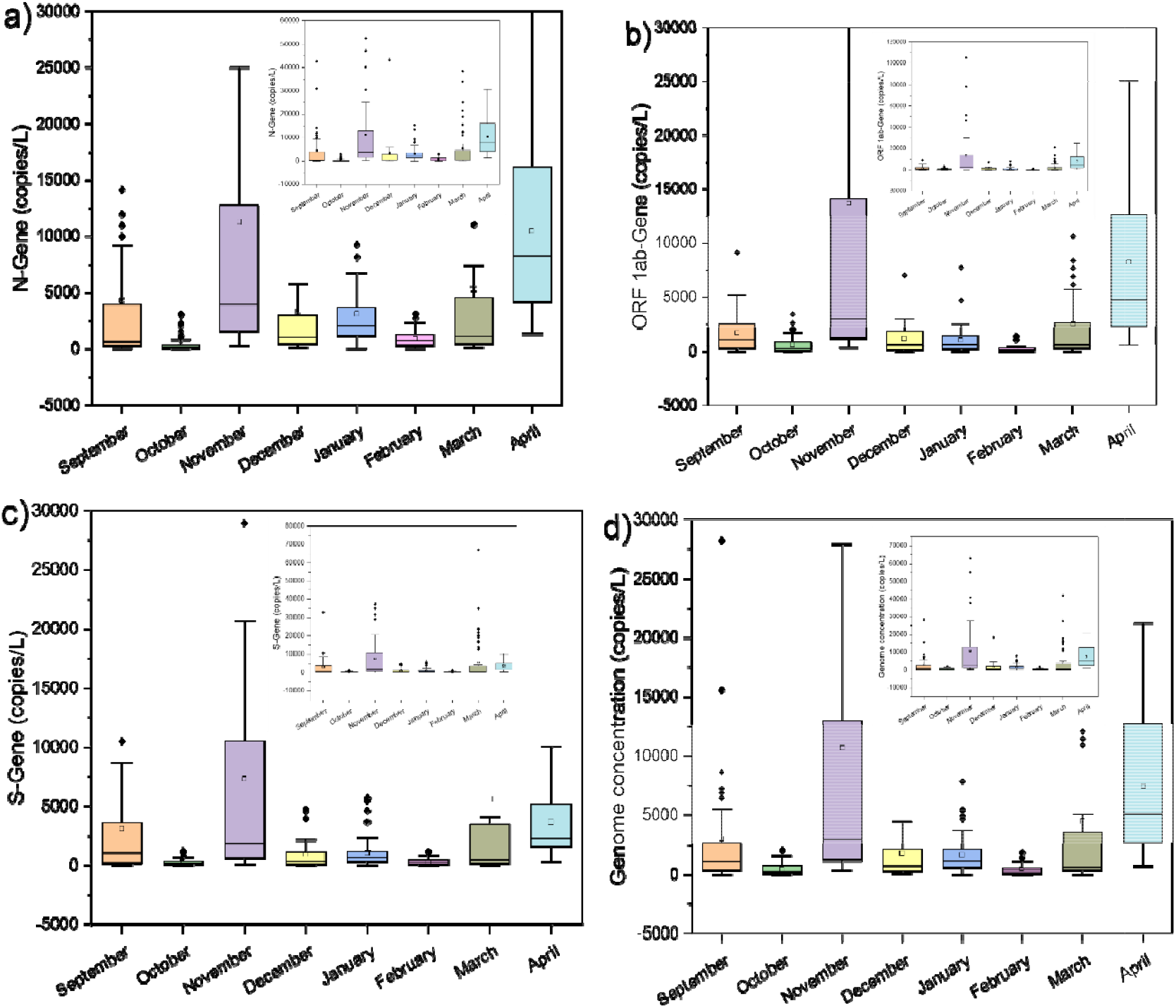
Monthly variations in average SARS-CoV-2 gene copies collected from different STPs in Ahmedabad, a) N-Gene, b) ORF1 ab Gene, c) S-Gene, and d) Genome concentration.

There had been a decrease of 20.47% in active cases in October 2020 with respect to September, followed by a rise of 1.82% in November 2020 compared to the preceding month, October. The rise of active cases of November with respect to October is analogous to a change of 59 cases (3,234 cases on 1^st^ November – 3,293 on 26^th^ November). Though the percentage change in active cases appears to be insignificant, the sharp increase in SARS-CoV-2 gene concentration can be attributed to a huge 37.93% increase in average daily confirmed cases in November 2020 (i.e., 252 cases) as compared to October 2020. (i.e., 183 cases). The casual and reluctant attitude of people during the festive season in India (mid-Oct to mid-Nov) might be the reason for the surge in COVID-19 cases. This was followed by a decline of 2.27% in active cases in December with respect to November (2998 cases on 14^th^ December 2020 – 2930 cases on 28^th^ December, 2020). In January 2021, there was a further decrease of 49% in the number of active cases with respect to December 2020 (2894 cases on 4^th^ January 2021 – 1478 cases on 29^th^ January 2021). Likewise, in February 2021, there was a reduction of 36% in active cases with respect to January 2021 (855 cases on 8^th^ February 2021 – 548 cases on 22^nd^ February 2021). Finally, a 3-fold increase in the number of active cases was noticed in the month of March with respect to February. This corresponds to a change of 1455 cases (642 cases on 1^st^ March, 2022 -2097 cases on 30^th^ March, 2022). Lockdown was imposed upon the city in April 2021, when the number of active cases had already reached 7165 (as on 12^th^ April, 2021). A whopping rise in the average daily confirmed cases in March 2021 (i.e., 317 cases) and April 2021 (i.e., 1017 cases till 12^th^ April) was noticed due to the second wave of COVID-19.

One of the main purposes behind WBE for COVID-19 detection is its capability to induce an early warning of the outbreak of disease or emergence of new trends within communities. The distinguishable increase in the number of active cases viz. 7165 appeared on 12^th^ April 2021, which is 2 weeks post the significant increase in the viral genome concentration in wastewater samples (on 30^th^ March 2021) (**Fig. 2**). Therefore, this time period of 2 weeks could be adequately utilized to control the pandemic situation in the city. Some other studies around the globe reported early detection of SARS-CoV-2 RNA in wastewater even before the first report of clinical diagnosis (**Table 2**). For example, in Netherlands, SARS-CoV-2 genetic material was detected in wastewater in February, even before the official declaration of the first case (Madema et al., 2020). Similarly, La Rosa et al. (2020) reported SARS-CoV-2 genetic material in wastewater samples from two different cities in Italy before the first official documented report. Likewise, Randazzo et al. (2020) identified SARS-CoV-2 RNA in wastewater samples from Spain. Thereafter, various studies have been carried out where the presence of SARS-CoV-2 RNA in wastewater samples were detected and reported successfully (Ahmed et al., 2020, Kumar et al. 2020 a,b). The researchers from Gujarat found the genomic traces of the B.1.617.2 in wastewater samples before a month of a clinically confirmed case of the same variant in Ahmedabad, Gujarat (Joshi et al., 2021). A few studies, however, focused on evaluating the potential of WBE on the temporal scale with respect to the changes in COVID cases.

**Fig. 2.**
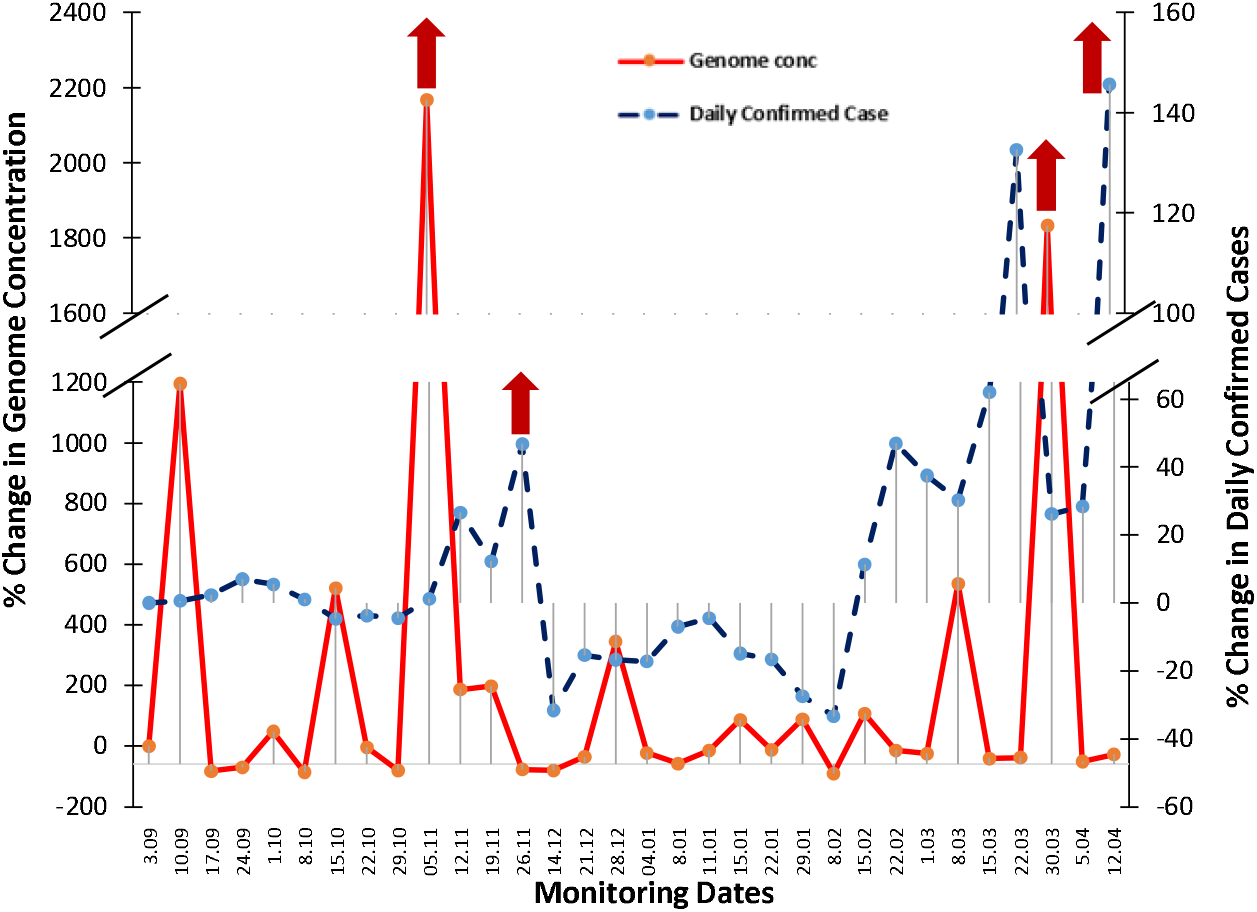
Potential and evidence of wastewater-based epidemiology surveillance of Covid-19 pandemic as an early warning tool in Ahmedabad

**Table 2.** Country-wise detection of SARS-Cov-2 viral RNA in wastewater before the confirmed COVID-19 cases and related casualty data

| **Name of the country** | **Confirmed presence in wastewater** | **First reported cases** | **Date of National Lockdown Measures** | **National casualties** | **Reference** |
| --- | --- | --- | --- | --- | --- |
| Amsterdam (New Zealand) | Feb 6, 2020 | Feb 27, 2020 | Mar 15, 2020 | 3134 | Medema et al., 2020 |
| Barcelona (Spain) | Jan 25, 2019 | Feb 25, 2020 | Mar 14, 2020 | 18276 | Vallejo et al., 2020 |
| Milan (Italy) | Dec 18, 2019 | Feb 21, 2020 | Mar 9, 2020 | 31106 | Rosa et al., 2020 |
| Brisbane South (Australia) | Feb 27, 2020 | Mar 13, 2020 | Apr 3, 2020 | 2121 | Ahmed et al., 2020 |
| Southeast England | Feb 11, 2020 | Feb 13, 2020 | Mar 23, 2020 | 1800 | Martin et al., 2020 |
| Ishikawa (Japan) | Feb 19, 2020 | Feb 21, 2020 | July 29, 2020 | 316 | Hata et al., 2020 |
| Toyama (Japan) | Mar 30, 2020 | Apr 4, 2020 | July 29, 2020 | 236 | Hata et al., 2020 |
| Santa Catarina, (Brazil) | Nov 27, 2019 | Mar 03, 2020 | Mar 21, 2020 | 51 | Fongaro et al., 2020 |

### 3.2. Early warning capability

The current investigation is based on our first proof of concept study, in which we discovered SARS-CoV-2 genetic material in wastewater and asserted that it may be used for community COVID-19 monitoring (Kumar et al. 2020a). The percentage change in genome concentration level on a particular date maintained a positive correlation with the confirmed cases registered 1-2 weeks later by the regulatory authority based on clinical tests (**Fig. 2**). This authenticated the early warning capability of WBE for COVID-19 monitoring on a temporal scale. However, no linear relationship exists between the SARS-CoV-2 gene concentration and epidemiological data. Therefore, we demonstrated the relationship between percentage changes in SARS-CoV-2 genome concentration and confirmed cases which can be used as a pre-alarming tool, as it offers a lead of around 2 weeks for the upcoming scenario. From the present study, we can see that on 8^th^ October, 2020, a sharp decline of 134% was noticed in the percentage change in the genome concentration which was followed by c.a. 4.8% decline in the percentage change in daily confirmed COVID cases on 22^nd^ October, 2020. Similarly, on 5^th^ November, 2020, a significant increase of >22-folds was noticed in the in the genome concentration compared to the earlier sampling date, which was followed by 11.16 and 45.58% increment in the percentage change in confirmed COVID cases on 19^th^ November and 26^th^ November, 2020, respectively. In contrast, more than >1,000% and 500% increase were observed in percentage change in SARS-CoV-2 effective gene concentration in wastewater in early September and mid-October, respectively. However, no notable increase in the number of confirmed cases was detected 1-2 weeks later. In spite of this, the aforementioned technique exhibited positive prediction in most of the cases during the study period. On 4^th^ January, 2021 a sharp decline of 366% was noticed in the percentage change in the genome concentration which was followed by 12.8% decline in the percentage change in confirmed COVID cases on January 11^th^, 2021.

The variations in SARS-CoV-2 RNA in wastewater was further detected and quantified to understand the pandemic situation during the second wave in Ahmedabad. On March 30th, 2021, a steep hike of >1800% in the percentage change in the genome concentration was noticed compared to the earlier sampling date, which was followed by 120% increment in the percentage change in confirmed COVID cases on April 12th, 2021 (**Fig. 2**). Therefore, the severity of the pandemic situation can be predicted 1-2 weeks prior to the official reports on the basis of clinical tests. The results from the study highlighted the potential of WBE monitoring as an early warning tool for COVID-19 in the presence of adequate SARS-CoV-2 genetic material in wastewater samples. The findings of the study further supported those of Ahmed et al. (2020b), who detected a longitudinal decline in the presence of SARS-CoV-2 RNA with the subsidence of the first epidemic wave.

These findings were further supported by Spearman’s rank correlation for the early warning potential of WBE. The results showed the highest rank correlation coefficient was observed in the ORF1 ab followed by S and N genes for all pumping stations (PS) and sewage treatment plants (STP). In the three PS, Santivan PS, Paldi PS and Ranip PS, the time lag was negative for all viral genes, and for the ORF1ab gene, which had the highest rank correlation coefficient, the time lag was -8 days for Santivan PS (r=0.45, p<0.01), -10 days for Paldi PS (r=0.54, p<0.01), and -11 days for Ranip PS (r=0.60, p<0.01) (Fig. 3). These PS showed that we could detect trends in viral infection status from the wastewater approximately 10 days earlier than the clinical examination. Although the rank correlation coefficients were smaller than those in the ORF1 ab gene, the same three PS also showed negative time lags in the N and S genes at about the same time as in the ORF1 ab gene. In Motera PS, the time lag was negative for all the genetic regions except ORF. Among N, S, and average genome regions, S showed the highest rank correlation coefficient (r=0.48, p<0.01) with a time lag of -8 days (Fig. 3). Likewise, In Vatva PS, negative time lag was noticed for all genes except N. The highest rank correlation coefficient was observed for ORF1 ab gene (r=0.38, p<0.02) with a lag time of -16 days. However, in the case of STP Vinzol, Odhav, Satyam, and Maninagar PS, no negative lead time was noticed for all the genes. The latter can be ascribed to i) issue with the selection of sampling point/ representative sample; ii) disparity in the actual infected individuals and clinical data. Nevertheless, following observations were critical in Spearman’s rank correlation between new daily positive cases and Ct values of different qPCR detection genes of different sites, i) ORF1 ab gene can be used as a marker gene for the early detection or changes in the spike of COVID-19 cases, and ii) WBE can be used for the early detection of COVID-19 at sub-city levels reflected by a clear-cut lead time. Therefore, it is suggested that we would be able to predictably capture the signs of the trend of increase or decrease in the number of infected people from wastewater.

**Fig. 3.**
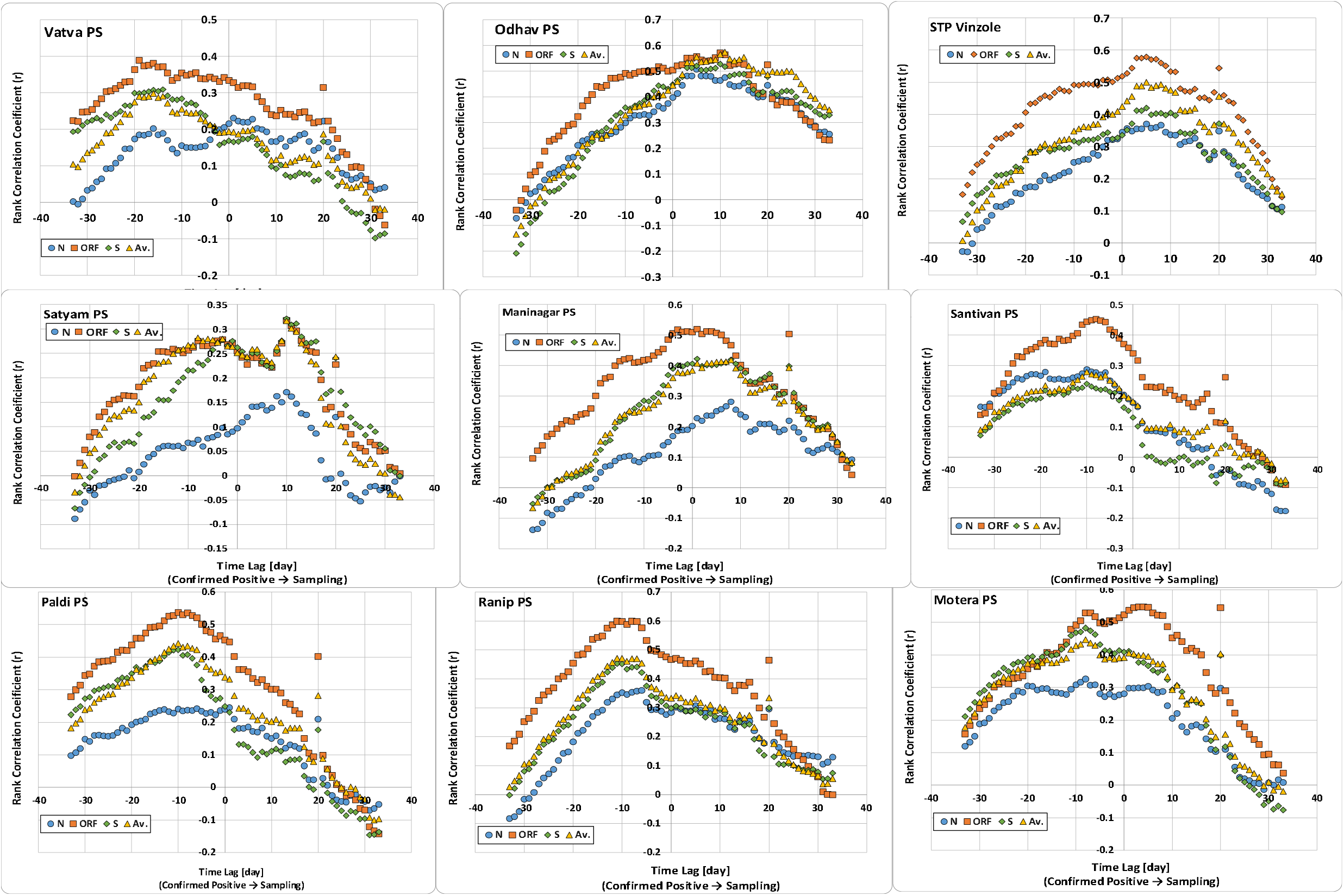
Rank correlation coefficient with the concentration of the SARS-CoV-2 gene in wastewater.

**Fig. 4.**
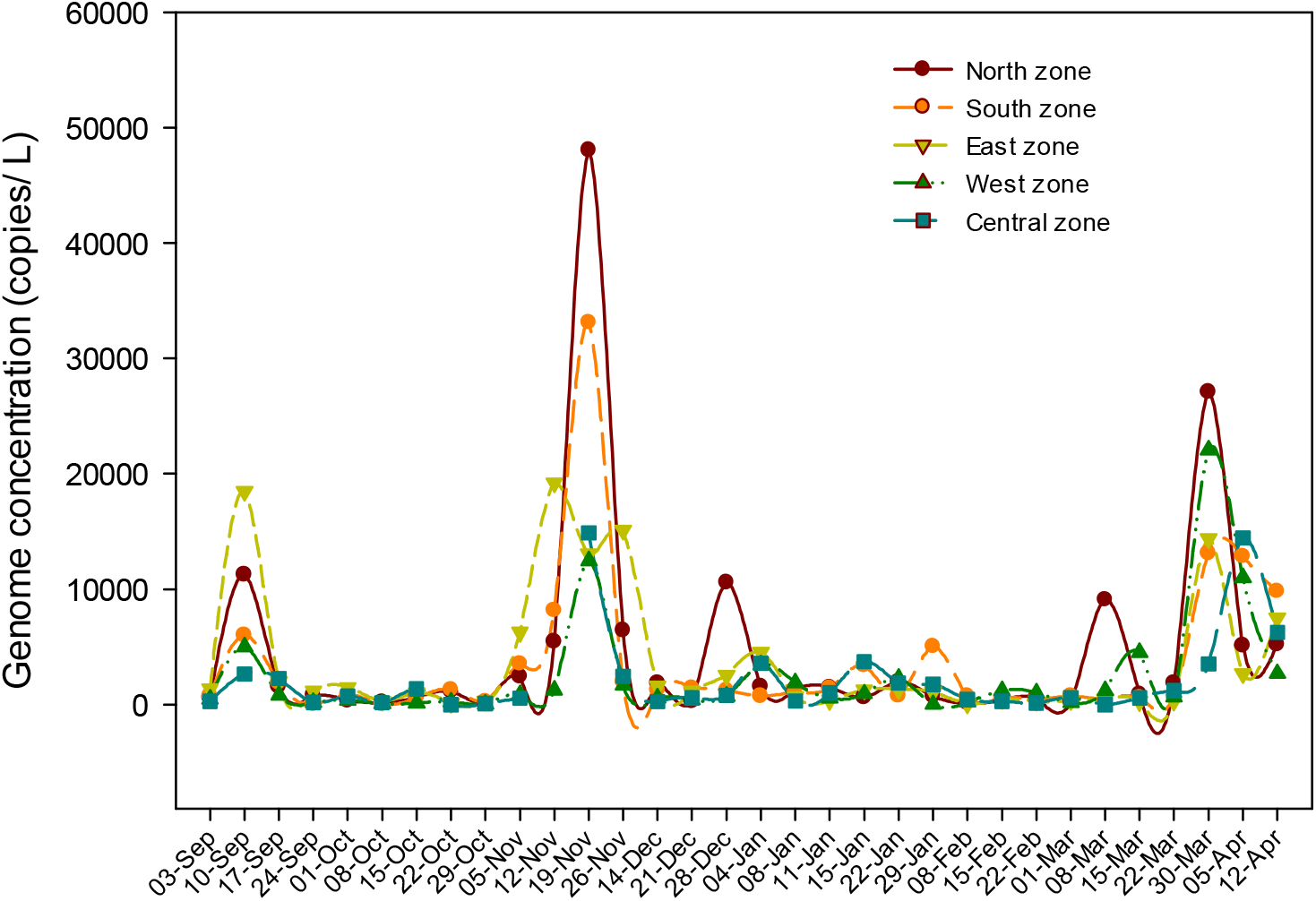
Zone-wise COVID-19 pandemic status in Ahmedabad city.

On comparing the SARS-CoV-2 genome concentration in wastewater of Ahmedabad, we found high genome concentration during the first wave (November 2020) compared to the second wave (April 2021), while daily new confirmed cases were much higher in the second wave in comparison to the first wave. The latter can be ascribed to i) the greater infectivity and transmissibility of the Delta variant (B.1.617.2) compared to the Alpha variant (B.1.1.7), which led to increased clinical testing during the second wave; ii) more asymptomatic patients and less clinical testing during the first wave.

### 3.3. SWEEP-based city zonation and identification of hotspots

On the basis of SARS-CoV-2 genome concentration in wastewater samples, we successfully recognized locations that are highly susceptible for COVD-19 infection and its transmission among the community. Despite not having explicit epidemiological data at the ward level/sampling locations, the variations of SARS-CoV-2 gene concentration in wastewater samples were sufficient to classify the city, despite not having explicit epidemiological data at the ward level/ sampling locations. In September, 2020 maximum effective gene concentration was displayed by the wastewater samples collected from the east zone (5.7×103 copies/ L), followed by the north zone (3.5×10^3^ copies/ L) (**Fig. 3**). Likewise, in November, 2020 the north (Motera and Ranip) and east (Odhav and Satyam) zones were particularly affected, with an average genome concentration of 1.6×104 and 1.3×10^4^ copies/L, respectively (**Fig. 3**). Even though areas present in the north and east zones revealed high virus genetic load, a sharp rise in SARS-CoV-2 RNA could be seen in all the zones in November 2020. On 28^th^ December, 2020 the north zone showed higher effective gene concentration (1.1× 10^4^ copies/L) as compared to the other zones. On 8^th^ March, 2021 an ascent in virus genetic load (9.1×10^3^ copies/L) could be seen from the north zone. At the end of March, 2021 (30^th^ March, 2021), the wastewater samples collected from the north zone showed maximum effective gene concentration (2.7×10^4^ copies/L), followed by the west zone (2.2×10^4^ copies/L), even though sharp rise in SARS-CoV-2 RNA was noticed in all the zones.

This implies the capability of SWEEP technology to distinguish the study area at the sub-city or zone level based on SARS-CoV-2 gene concentration. SWEEP data can offer insight into the actual extent of the infection due to the SARS-CoV-2 since it covers both asymptomatic and symptomatic patients. It is, thus, possible to identify hot spots within the city, which can assist in increasing the preparedness in advance. Contrarily, clinical surveillance usually falls short while classifying the city into distinct zones as it is primarily dependent upon the location of test centres. Also, it does not take the number of asymptomatic patients into account.

## 4. Conclusions

Wastewater-based epidemiology holds a lot of promise as a favourable tool that could be used to detect real-time and early disease signals. It could also be utilized to determine emerging hot spots in the monitoring of COVID-19 prevalence at the community level. WBE could prove to be extremely essential in an Indian content, where there is a scarcity of resource both in terms of disease management and diagnosis. In the present study, a temporal variation of the presence of SARS-CoV-2 RNA in wastewater was carried out for a period of eight months in Ahmedabad, India. The relation between the percentage change in effective gene concentration level and confirmed cases which followed a similar trend on the temporal scale, further validated the results of the study. Additionally, the findings of the study successfully detected the resurge of viral RNA load in wastewater, approximately 2 weeks prior to the second wave of COVID-19 in Ahmedabad, India. This gap of c.a. 2 weeks between the change in genome concentration in wastewater samples and the number of confirmed cases of COVID-19 unveils the potential of WBE monitoring as an early warning. Likewise, Spearman’s rank correlation suggested the highest rank correlation coefficient for ORF1 ab gene followed by S and N genes. Also, the results showed that we could detect trends in viral infection status from the wastewater approximately 10 days earlier than the clinical examination. This time gap may prove to be sufficient to take effective management interventions to stop the spread of the disease and further assist the authorities to identify the hotspots within a city. Additional research should be promoted to develop a predictive model that can interpret SWEEP data to policymakers to boost the awareness and management of pandemics.

## Data Availability

All data produced in the present study are available upon reasonable request to the authors

https://www.covid19india.org/state/GJ

## Notes

The authors declare no competing financial interest.

## Acknowledgement

This work is funded by UNICEF, Gujarat. We acknowledge the help received from GPCB and AMC. All data produced in the present study are available upon reasonable request to the authors

